# Seroprevalence of antibodies against SARS-CoV-2 among public community and health-care workers in Alzintan City of Libya

**DOI:** 10.1101/2020.05.25.20109470

**Authors:** Abdulwahab M. Kammon, Ali A. El-Arabi, Esadk A. Erhouma, Taha M. Mehemed, Othman A. Mohamed

## Abstract

A study was conducted to determine the seroprevalence of antibodies against SARS-CoV-2 among public community and health care workers in Alzintan City, Libya. During the period from 2/4/2020 to 18/5/2020, a total of 219 blood samples were collected and analyzed for the presence of antibodies against SARS-CoV-2. Collection of samples were divided in two categories; random samples from public community and samples from health care workers belonging to two Governmental hospitals and one private clinic. One Step Novel Coronavirus (COVID-19) IgM/IgG Antibody Test was used. Out of the 219 samples tested, 6 (2.74%) samples were seropositive for SARS-CoV-2. All health-care workers were tested negative. All positive cases were females and 5 of them aged between 44 to 75 years and one aged 32 years. The prevalence in young females (≤40 years) was 1.4% in total young females tested in the study and 1.75% in young females taken from public community. The prevalence in older females aged (≤ 40 years), was 11.1% in total females tested and 13.9% in females taken from public community. In conclusion, the preliminary investigation of SARS-CoV-2 revealed considerable prevalence in Alzintan City although the disease seems to be in its mild form. Active surveillance studies with high number of samples using both virological and serological tests are in urgent need.

## Introduction

Coronaviruses (CoVs) are a large family of viruses that are phenotypically and genotypically diverse. CoVs are enveloped viruses containing single-stranded positive-sense RNA that can cause illness in birds, mammals and humans. The viral genome is about 27-32 kb, which encodes for both structural and non-structural proteins. The coronavirus disease (COVID-19) is a novel viral disease reported firstly in a cluster of human pneumonia cases in December 2019 in Wuhan, China. The disease is caused by SARS-CoV-2 leading to pandemic spread throughout the world.

To date, in Libya a total of 3705 suspected samples were tested by the National Center of Disease Control (NCDC) for presence of SARS-CoV-2 in the whole country. 65 cases were tested positive with only 3 deaths, 35 recovered and active cases (still under treatment in the isolation wards). The first positive case was reported in Tripoli in 24/3/2020. All positive cases thus far were from the cities of Tripoli, Misrata, Benghazi and Surman. Three suspected cases from Alzintan city were previously submitted to NCDC and tested negative.

The main test used by NCDC is real time reverse transcriptase polymerase chain reaction (rRT-PCR). rRT-PCR is a quantitative analysis for SARS-CoV2 and considered as the gold standard for diagnosing COVID-19 [1]. In studies conducted in China, chest computed tomography (CT) scans were widely utilized as a diagnostic tool for COVID-19 Pneumonia [2,3]. Lung involvement can be detected in patients with COVID-19 on a CT scan in advance of the symptoms typical for pneumonia [4] and a positive result on rRT-PCR [1,5]. Accordingly, Chinese health authorities defined the suspected COVID-19 cases based on some criteria including epidemiological history, clinical signs, CT scans and CBC [6].

Although their clinical usefulness has yet to be thoroughly evaluated, an immunochromatographic (IC) assay for IgM and IgG antibodies against the virus is widely accepted as a point-of-care test because it is an easy-to-perform, rapid, and high-throughput method for diagnosing viral infections. Recently, several commercial IC assays that detect IgM/IgG antibodies against SARS-CoV2 have become available. Since the identification of asymptomatic patients with COVID-19 is important to prevent transmission of infection, and due to the small number of samples tested by NCDC using rRT-PCR in Libya, this study was conducted to evaluate the seroprevalence of antibodies against SARS CoV-2 among public community and health care workers in Alzintan City.

## Materials and Methods

### Collection of samples

During the period from 2/4/2020 to 18/5/2020, a total of 219 blood samples were collected and analyzed for the presence of antibodies against SARS-CoV-2. Collection of samples were divided in two categories; random samples from public community and samples from health-care workers belonging to two Governmental hospitals and one private clinic (Table 1). The random samples were collected from patients visiting Obstetrics and Gynecology Center and Blood Bank Center (Both are located in the same building) in Alzintan City for regular medical checkup, laboratory analysis, blood donation and for MRI. The study was conducted at the laboratory of Alzintan Blood Bank Center. Ethical approval was provided by Alzintan Municipality Committee for Combatting COVID-19 Pandemic.

**Table 1.** Collection of samples for the seroprevalence study of SARS-CoV-2 among public and health care workers in Alzintan City of Libya.

| Category | Location | Number of Samples |  |  |  | Total |
| --- | --- | --- | --- | --- | --- | --- |
|  |  | Female |  | Male |  |  |
|  |  | ≤40 Years | □40 Years | ≤40 Years | □40 Years |  |
| Health Care Workers | Obstetrics and Gynecology Center and Blood Bank Center | 10 | 4 | 7 | 1 | 22 |
|  | Alzintan Trauma and Surgery Teaching Hospital | 2 | 1 | 27 | 6 | 36 |
|  | Altaqwa Private Clinic | 2 | 4 | 11 | 2 | 19 |
| Random public | Random people from Alzintan city | 57 | 36 | 9 | 28 | 130 |
|  | International workers | 0 | 0 | 12 | 0 | 12 |
| Total |  | 71 | 45 | 66 | 37 | 219 |

### Detection of IgM and IgG antibodies for SARS-CoV2

Blood samples were collected in tubes with clot activator. Samples were then centrifuged at 4000 rpm for 10 minutes for separation of sera. IgM/IgG antibody tests for SARS-CoV-2 were performed using the One Step Novel Coronavirus (COVID-19) IgM/IgG Antibody Test (Lateral flow method) (Wondfo, Guangzhou, China) according to the manufacturer’s instructions. It is an immunochromatographic (IC) assay for rapid, qualitative detection of SARS-CoV-2. In brief, 10μL serum was added to the sample port (well) of the IC assay and subsequently, 2-3 drops of sample buffer were added to the buffer port (well), and the results were interpreted after a 15–20 min incubation. The presence of only the control line indicates a negative result and valid test. The presence of both the control line and the IgM or IgG antibody line indicates a positive result for IgM or IgG antibody, respectively.

## Results and Discussion

Seroprevalence of antibodies against SARS-CoV-2 in public community and health–care workers in Alzintan City of Libya is summarized in Table 2. The study included 219 cases (115 females and 104 males). Out of the 219 samples tested, 6 (2.74%) samples were seropositive for SARS-CoV-2 using IgM/IgG antibody rapid test. This result is similar to the seroprevalence of antibodies to SARS-CoV-2 in Santa Clara County, USA in early April which was 2.8% using lateral flow immunoassay (LFIA) [7]. A combined IgG/IgM test seems to be a better choice in terms of sensitivity than measuring either antibody type alone. Comparing ELISA to LFIA; both methods yielded high specificity reaching levels around 99%. ELISA performed better in terms of sensitivity (90-94%) followed by LFIA sensitivity ranging from 80% to 86% [8]. According to the manufacturer of the LFIA used in the current study, the sensitivity is 86.43% and the specificity is 99.57% using samples collected from 313 confirmed cases and 283 negative cases. The assay showed no cross reactivity with 17 viruses and mycoplasma including parainfluenza virus, influenza type A and B viruses and mycoplasma pneumonia. It is not clear, however, if cross reaction with other mild human non-SARS-CoV-2 strains, such as coronavirus HKU1, NL63, OC43, or 229E, could interfere with serological tests. Therefore, correlation with clinical feature is very important.

**Table 2.** Result of seroprevalence study of SARS-CoV-2 among public and health care workers in Alzintan City of Libya.

| Category | Location | Number and % of Positive Samples |  |  |  | Total |
| --- | --- | --- | --- | --- | --- | --- |
|  |  | Female |  | Male |  |  |
|  |  | ≤40 Years | □40 Years | ≤40 Years | □40 Years |  |
| Health Care Workers | Obstetrics and Gynecology Center and Blood Bank Center | 0/10 | 0/4 | 0/7 | 0/1 | 0/22 |
|  | Alzintan Trauma and Surgery Teaching Hospital | 0/2 | 0/1 | 0/27 | 0/6 | 0/36 |
|  | Altaqwa Private Clinic | 0/2 | 0/4 | 0/11 | 0/2 | 0/19 |
| Random public | Random people from Alzintan city | 1/57 (1.75%) | 5/36 (13.9%) | 0/9 | 0/28 | 6/130 (4.6%) |
|  | International workers | 0 | 0 | 0/12 | 0 | 0/12 |
| Total |  | 1/71 (1.4%) | 5/45 (11.1%) | 0/66 | 0/37 | 6/219 (2.74%) |

Seroprevalence studies can provide relevant information on the proportion of people who have experienced a recent or past infection. They are relevant when conducted in the community, but also for critical population subgroups such as nursing homes or health care facilities [9]. Moreover, knowledge of past infection among health-care workers could be useful for avoiding unnecessary quarantines and for health care resource planning [10]. In our study, health-care workers were tested negative. This may reflect the awareness of the health-care workers and the effectiveness of the protection measures taken while dealing with patients. All 6 positive cases were females, 5 of them aged between 44 to 75 years and one aged 32 years. The prevalence in young females (≤40 years) was 1.4% in total young females tested in the study and 1.75% in young females taken from public community. The prevalence in older females aged (≥40 years), was 11.1% in total older females tested and 13.9% in older females taken from public community. This is consistent with the fact that people with low immune function particularly old aged people are at high risk of infection [11]. In contrast to our findings, many other studies reported higher prevalence in males compare to females [11,12]. Another study reported similar susceptibility to SARS-CoV-2 between males and females in 1,019 patients who survived the disease [13]. Therefore, more studies may be required to consider the gender as a risk factor for COVID-19. All positive cases found by our study were belonging to public community. One of the positive cases reported with mild respiratory symptoms and had a chest CT scan showing the typical findings of COVID-19 pneumonia in the form of lower lung lobe subpleural mixed ground glass opacities and consolidations. The patient has recovered with antibiotics. The test was repeated for her after one month and the result was positive with no respiratory complications. Surprisingly, her husband tested negative for antibodies against SARS-CoV-2, suggesting that transmission of the virus between individuals may not be as straight forward as was indicated by normal human interaction.

Interestingly, the first case confirmed positive in this study was tested on April 8, 2020, which is only 15 days after the first case was confirmed positive by NCDC on March 24, 2020 using rRT-PCR. This indicates that the virus has been widely spread among Libyan population way long before it was first detected by NCDC considering that the method of identification used by NCDC was to detect viral antigen, while our method was to detect antibodies against the virus, which naturally results after viral infection has occurred and lasts for months afterward.

The high prevalence of COVID-19 (2.74%) identified by our study suggests that the viral infection amongst Libyan population is far higher than that identified by NCDC. If we were to apply the same analogy inferred by this study on the ~ 7 million Libyan population size, we would expect a close to 200,000 people infected by the virus, which is almost 3,000 times higher than the number of positive cases identified by NCDC.

In conclusion, the preliminary investigation of SARS-CoV-2 revealed considerable prevalence in Alzintan City although the disease seems to be in its mild form. It is elusive that relatives of some COVID-19 rRT-PCR confirmed patients were negative in the cities of Tripoli, Misrata and Surman. Similarly, in the city of Alzintan, relatives of some confirmed positive patients have tested negative suggesting more complicated form of viral transmission than has previously outlined. Therefore, isolation, sequencing and phylogenetic analysis of SARS-CoV-2 circulating in Libya is strongly suggested. A vast active surveillance studies with high number of samples using both virological and serological tests are in urgent need although the presence of hard challenges facing the health care workers due to the civil war, insecurity and lake of equipment and reagents.

## Data Availability

None

## Acknowledgments

The authors acknowledge the Municipality of Alzintan for providing the rapid test. Special thanks to the Director of Alzintan Blood Bank Center and the staff for their assistance in samples collection and providing the lab for analysis. Thanks also go to all Health Care Institutions in Alzintan City for their collaboration and support.

